# AI-algorithm training and validation for endometrial CD138+ cells in infertility-associated conditions; polycystic ovary syndrome (PCOS) and recurrent implantation failure (RIF)

**DOI:** 10.1101/2023.12.05.23299423

**Authors:** Seungbaek Lee, Riikka K. Arffman, Elina K. Komsi, Outi Lindgren, Janette A. Kemppainen, Hanna Metsola, Anne Ahtikoski, Keiu Kask, Merli Saare, Andres Salumets, Terhi T. Piltonen

## Abstract

Immunohistochemical analysis of CD138+ plasma cells has been applied for detecting endometrial inflammation, especially chronic endometritis (CE). In this study, we developed for the first time an artificial intelligence (AI) algorithm, AITAH, to identify CD138+ plasma cells within endometrial tissue, focusing on two infertility-related conditions: polycystic ovary syndrome (PCOS) and recurrent implantation failure (RIF). We obtained 193 endometrial tissues from healthy controls (n=73), women with PCOS (n=91), and RIF patients (n=29) and compared CD138+ cell percentages across cycle phases, ovulation status, and endometrial receptivity. We trained AITAH with CD138 stained tissue images, and experienced pathologists validated the training and performance of AITAH. AITAH, with high accuracy in detecting CD138+ cells (88.57%), revealed higher CD138+ cell percentages in the proliferative phase than in the secretory phase or in the anovulatory PCOS endometrium, irrespective of PCOS diagnosis. Interestingly, CD138+ percentages differed according to PCOS phenotype in the proliferative phase (p=0.01). Different receptivity statuses had no impact on the cell percentages in RIF samples. In summary, the AI-enabled analysis is a rapid and accurate tool to examine endometrial tissues, potentially aiding clinical decision-making. Here, the AI analysis demonstrated cycle-phase differences in CD138+ aggregations pattern, but no major alterations in PCOS or RIF samples.

## Introduction

Menstrual cycle-related fluctuations in sex hormones not only contribute to the growth and degeneration of the endometrial lining but also modulate the local immune environment by regulating innate/adaptive immune responses and endometrial immune cell populations: B and T lymphocytes, uterine natural killer (uNK) cells, plasma cells, macrophages, and dendritic cells^1–3^. As a sum effect, cellular immunity in the endometrium is high in the proliferative phase but declines toward the mid-secretory phase, facilitating successful implantation of the semi-allogenic embryo^2^.

Endometrial plasma cells can either be recruited from the circulation or developed locally from antigen-committed B cells^4^. CD138, a heparan sulfate proteoglycan (HSG) on the surface of plasma cells, serves as a receptor for growth factors and immune mediators, attracting plasma cells to the endometrium^5^. Endometrial CD138+ plasma cells have been used as a diagnostic biomarker for endometrial inflammation, including chronic endometritis (CE)^6–8^, and the number of CD138+ plasma cells is correlated with the severity of endometrial inflammation in women with reproductive failure^7–9^. Identifying isolated CD138+ plasma cell aggregates without other histological features of CE, such as stromal edema and increased stromal cell density, may also detect cases with potentially milder inflammation and related endometrial dysfunction^10^. However, current quantification methods of CD138+ cells are typically based on laborious and time-consuming microscopic assessments of only a few random areas from a sample^10^. These methods have limitations in accurately representing the entire slide and are susceptible to significant biases arising from both intra- and inter-observer variations^10–12^.

Convolutional neural networks (CNNs) are widely used to analyze histological patterns^13^. In particular, the artificial intelligence (AI) embodied by CNNs enables rapid analysis of whole tissue slides with high resolution, addressing the current limitations of manual histopathological performance^13^. To date, this promising and potent AI technique has been increasingly applied in various clinical research fields to predict the onset and progression of specific diseases^14–18^. Given that an imbalanced inflammatory milieu has been linked to infertility-associated conditions, such as polycystic ovary syndrome (PCOS)^19^ and recurrent implantation failure (RIF)^9,10^, this study utilized an AI algorithm to assess endometrial CD138+ cells in these two conditions.

PCOS is a common endocrine disorder characterized by hyperandrogenism (HA), systemic low-grade inflammation, insulin resistance, and anovulation^20^. The PCOS endometrium exhibits dysregulated immune profiles^19^ and altered receptivity-related proteomic profiles^19,21^, which could lead to poor reproductive outcomes^19,21^. However, many studies lack sufficient endometrial sample dating, often overlooking secretory phase samples despite occasional spontaneous ovulations and pregnancies in PCOS women. Moreover, although the presence of CD138+ cells varies depending on the menstrual cycle phase^8,12^, CD138+-based evaluation in PCOS endometrium across different cycle phases is lacking.

RIF, often defined as three or more failed *in vitro* fertilization (IVF) attempts with good-quality embryos transferred, significantly impacts IVF success rates^22^. Factors from the maternal side, including age, body mass index (BMI), and immunological factors^22^, as well as unprepared endometrium for embryo implantation (e.g., shifted window of implantation, impaired decidualization in stroma)^23,24^, have an impact on the implantation rate and reproductive outcomes^22,25^. Moreover, CE is prevalent in RIF patients, affecting up to 14% of this population^9^. Given the multifaceted underlying mechanisms of RIF, it is important to understand the association between endometrial receptivity and inflammatory processes, particularly CD138+ cell clustering in fully receptive samples without prominent CE.

In this study, we developed an AI algorithm, “Artificial Intelligence for Targeted Analysis of Histology” (AITAH), to detect stromal CD138+ cells, an indicator of the endometrial immune milieu. As a novel setup, we investigated the CD138+ cell occurrence to identify factors influencing the aggregation of CD138+ cells in relation to menstrual cycle phases and ovulatory status under PCOS conditions, as well as endometrial receptivity under RIF conditions.

## Materials and methods

### Tissue collection

#### PCOS and control samples

A total of 164 endometrial biopsy samples from 44 healthy controls and 61 women with PCOS were collected at the Oulu University Hospital (Oulu, Finland) from January 2017 to March 2020 (**Fig 1**, **Table 1**). The study was approved by The Regional Ethics Committee of the Northern Ostrobothnia Hospital District, Finland (65/2017), and informed consent was signed by all study subjects. All methods were carried out in accordance with relevant guidelines/regulations in line with the Declaration of Helsinki. All women were non-smokers and had not used any hormonal medication for at least three months prior to tissue collection. Some women gave samples in multiple phases in different cycles (only one biopsy per cycle), with a maximum of three samples per individual (depicted by multiple sample providers in **Fig 1**). Control women were healthy, had regular cycles, and were without PCOS symptoms. As per the international Guideline, PCOS was diagnosed according to the Rotterdam consensus, requiring the presence of at least two of the following clinical features: oligo-anovulation (OA), HA, and polycystic ovarian morphology (PCOM) ^26^. Some women with PCOS had occasional ovulations that were traced for secretory phase sampling and analysis. As for PCOS phenotype sub-analysis, PCOS women were divided based on their phenotype; 35 women with phenotype A (PCOM+OA+HA, most severe PCOS phenotype, metabolic) and 19 with phenotype D (PCOM+OA, mildest phenotype, reproductive), were identified (**Supplemental Table S1**)^26^. Endometrial biopsies were obtained by a suction curette (Pipelle), fixed in 10% formaldehyde for hematoxylin and eosin (H&E) staining and immunohistochemistry (IHC)^27^. The samples for ovulatory cycles were obtained in either the proliferative phase (PE) (cycle days 6–8) or the secretory phase (SE) on specific days after the luteinizing hormone (LH) surge, at +2–4 days (early secretory phase (ESE)), +7–8 days (mid-secretory phase (MSE)), or +11–12 days (late secretory phase (LSE)) (PE; 12 control, 24 PCOS, ESE; 15 control, 15 PCOS, MSE; 26 control, 21 PCOS, LSE; 20 control, 18 PCOS). The LH surge was detected using a Clearblue digital urine test in the morning, and the presence of the corpus luteum was confirmed by transvaginal ultrasonography (TVUS) via Voluson E8 (GE Healthcare Technologies, United States). The cycle phases were histologically determined from the H&E-stained slides by an experienced gyneco-pathologist. In the case of anovulation confirmed by clinical evaluation and histological analysis, 13 biopsies were obtained on any day convenient for the PCOS subjects.

**Figure 1.**
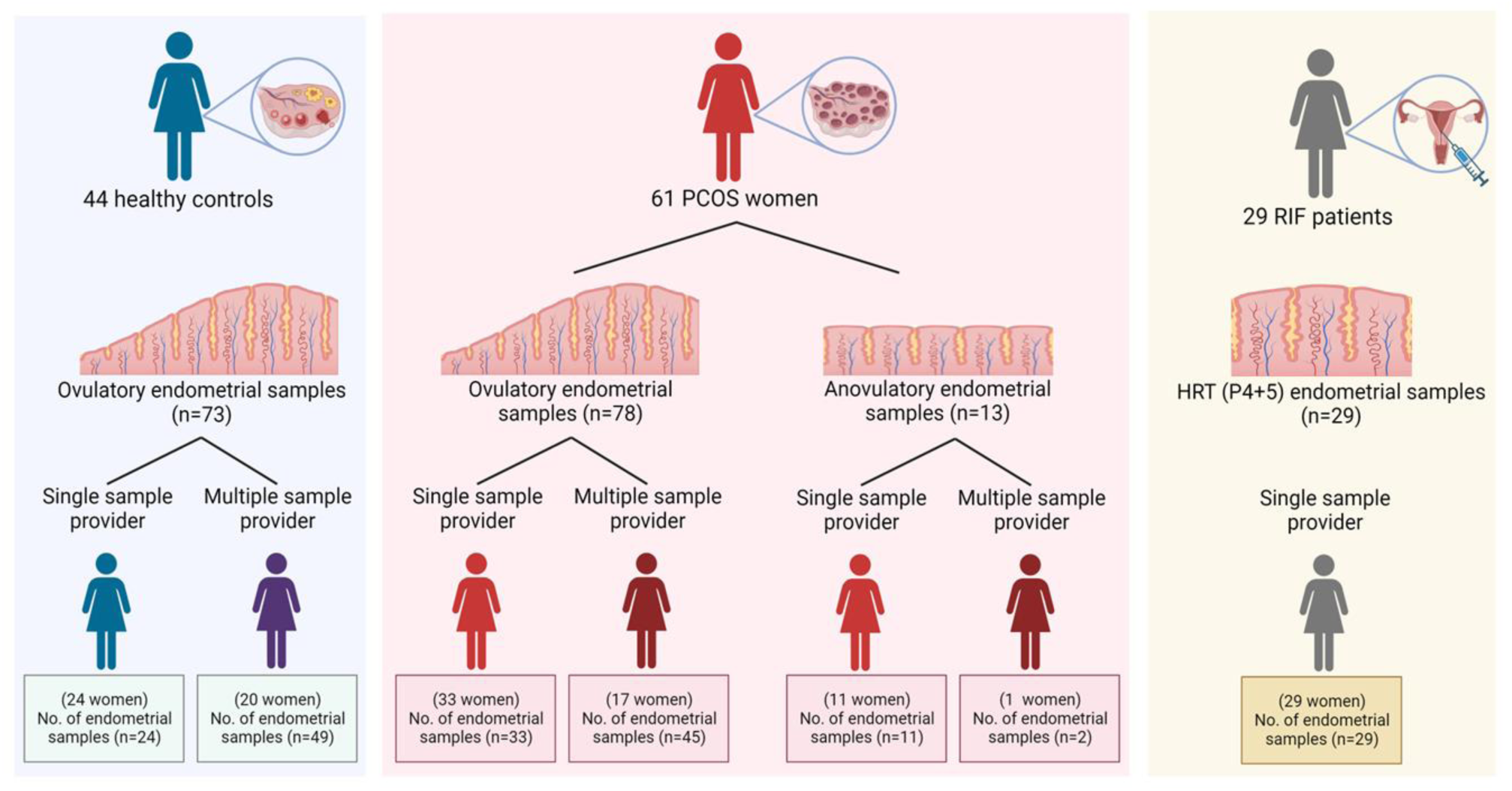
**The study subjects in the control and PCOS population** A total of 164 endometrial biopsy samples were collected from 44 healthy controls and 61 women with PCOS. The subjects were categorized based on whether they provided a single or multiple samples. In the case of women with PCOS, the subjects were further classified depending on their ovulatory status. A total of 29 samples were collected from 29 RIF patients who underwent IVF treatment. HRT (hormone replacement treatment), P4 (progesterone)

**Table 1.**
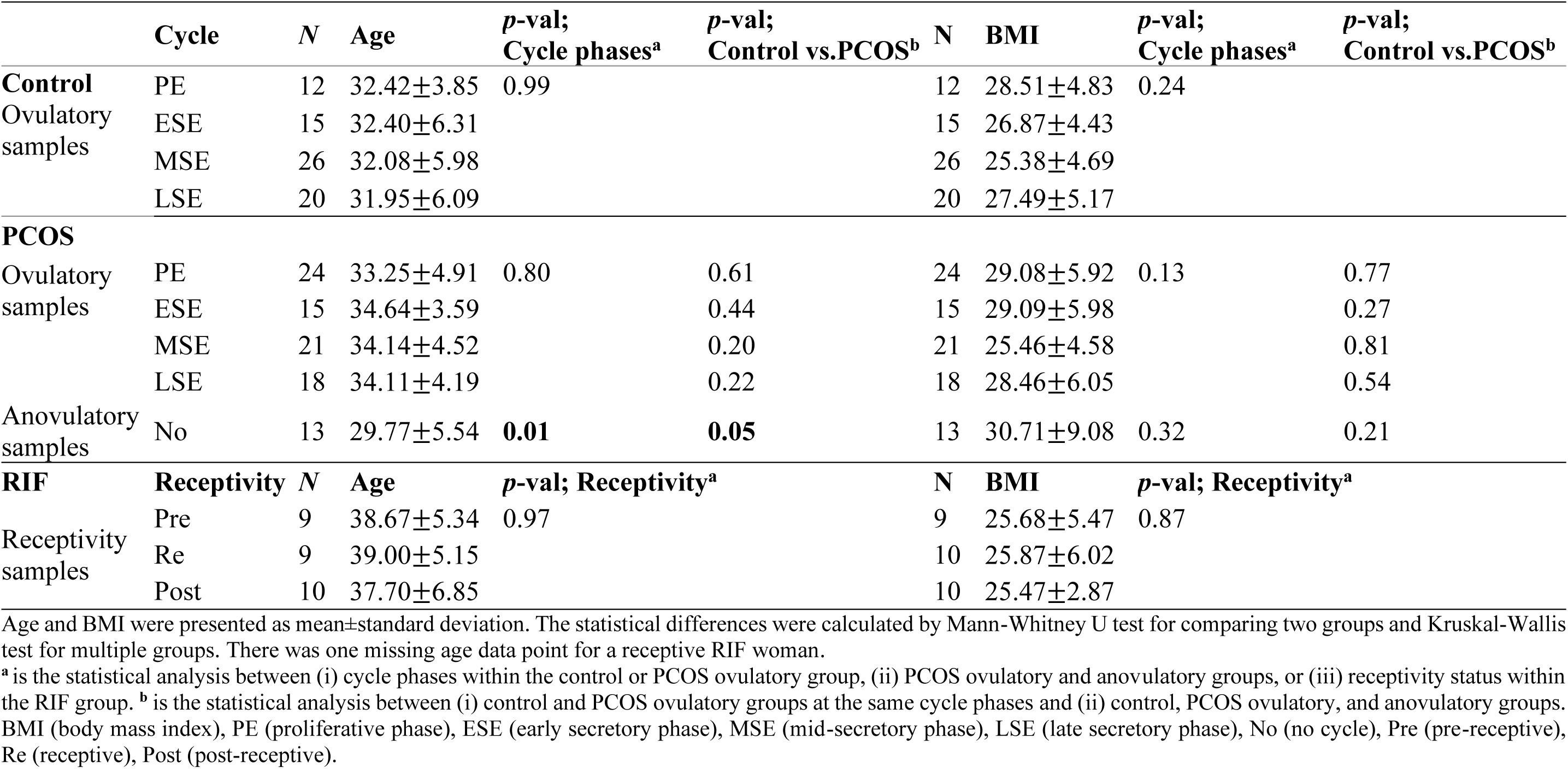
Subject and sample information of age and BMI in different cycle phases and receptivity status.

Endometrial thickness was measured by TVUS. Serum levels of anti-Müllerian hormone (AMH), LH, follicle-stimulating hormone (FSH), and sex hormone-binding globulin (SHBG) were measured with Elecsys assays (Roche) using a cobas e411 analyzer. Serum testosterone and progesterone (P4) levels were measured at the University of Eastern Finland using an Agilent 1290 Rapid Resolution LC System (Agilent, San Jose, CA, United States)^28^. Free androgen index (FAI) was calculated using the formula: testosterone (nmol/L) / SHBG (nmol/L) *100.

#### RIF samples

A total of 29 RIF samples were obtained from the beREADY endometrial receptivity testing laboratory (CCHT, Tartu, Estonia) from women who had undergone an average of 3.9 unsuccessful IVF cycles despite having good-quality embryos transferred (**Fig 1**). The study was approved by the Research Ethics Committee of the University of Tartu, Estonia (340T-12). The requirement for informed consent for obtaining RIF samples was waived by the same Research Ethics Committee as data was anonymized. All experiments were performed in accordance with relevant guidelines/regulations in line with the Declaration of Helsinki. Their anonymized data and endometrial biopsies were obtained prior to the subsequent IVF treatment^23^ using a Pipelle during a hormone replacement treatment (HRT) cycle on day 5 after initiation of P4 administration (HRT: P+5). The first day of P4 administration was considered day zero (HRT: P+0). All biopsies were placed into RNAlater (Ambion, United States), and then stored at -80℃ before use

### Endometrial receptivity testing

Gene expression profiling was conducted using the beREADY test at CCHT, involving 57 known endometrial receptivity genes along with four housekeeping genes (*SDHA*, *CYC1*, *TBP*, and *HMBS*)^23,24^. Endometrial RNA from RIF patients (n=29) was extracted using the Qiagen miRNeasy Mini kit, and according to the test results, the RIF samples were divided into pre-receptive (*n*=9), receptive (*n*=10), and post-receptive (*n*=10) (**Table 1**).

### Immunohistochemistry

The endometrial biopsies were processed accordingly: 5-µm paraffin-embedded tissue sections of controls (n=73) and women with PCOS (n=91) and 2.5-µm paraffin sections of RIF patients (n=29) were de-paraffinized in xylene and rehydrated in different grades of alcohol. Next, antigen retrieval with Tris-EDTA (pH 9) was performed in a microwave oven (800W) for 2 min. Dako peroxidase blocking solution (Dako S2023) was used to neutralize endogenous peroxidase for 5 min. The sections were incubated with 40x diluted mouse anti-human monoclonal antibody CD138 (MS-1793-S; Thermo Fisher Scientific) for 30 min at room temperature and then for 30 min with Dako EnVision polymer (Dako K5007), followed by a DAB working solution for 3 min. H&E staining was performed for 15 sec. The IHC staining for both controls and PCOS samples was performed at the University of Oulu (Oulu, Finland), and the staining for the RIF samples was processed at the Tartu University Hospital Pathology Department and the University of Tartu (Tartu, Estonia). Positive and negative controls from four different human tissues (i.e., tonsils, liver, pancreas, and appendix) were used for the CD138 antibody.

All IHC slides were scanned using a Leica SCN 400 Slide Scanner (Leica, Biosystems, United States), and the digitalized whole slide images (WSIs) were uploaded to Aiforia cloud (Aiforia Technologies Oy, Helsinki, Finland). Eleven slides with inadequate quality were excluded.

### AITAH algorithm training

Our previous AI algorithm, AINO, was modified and trained to detect CD138+ plasma cells in the endometrial stroma^16^. Our current AI algorithm, AITAH, was trained using morphological features derived from 150 WSIs across two layers: the regional layer and the object layer. All training was performed on regions of interest (ROI) drawn to remove noise and background interference that could affect the analysis. The regional layers were trained to segment the epithelium and stroma, followed by training the object layers to differentiate CD138- and CD138+ cells within the stroma. The training set consisted of a 28,363 mm^2^ region and 7,345 object layers (6,942 CD138-cells and 403 CD138+ cells). The algorithmic structure of AITAH and the training process are shown in **Fig 2a-d**.

**Figure 2.**
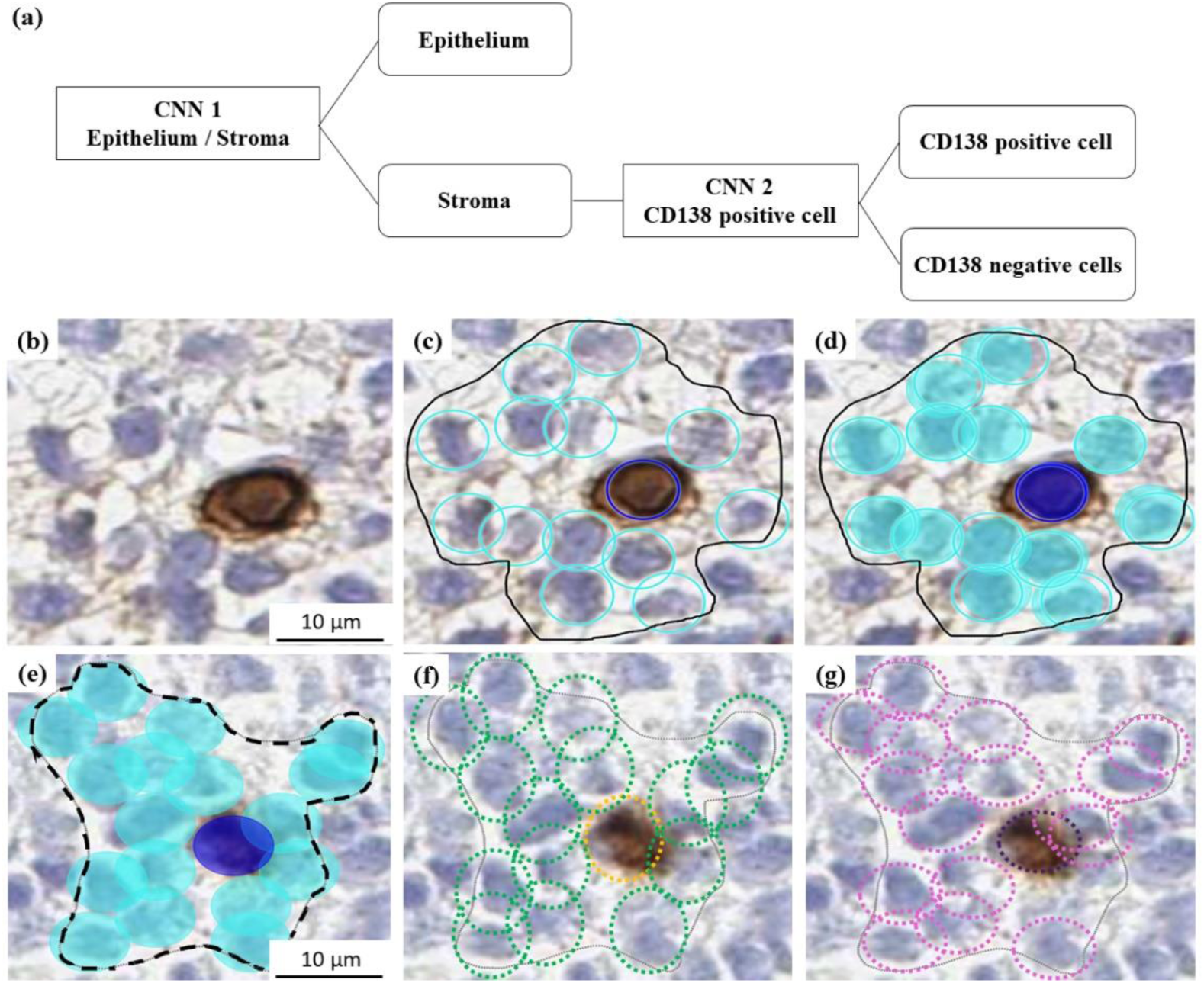
**Schematic overview of the AITAH algorithm and examples of training and validation.** (a) Structure of the convolutional neural networks (CNNs) model. CNN1 for the regional layer and CNN2 for the object layer. (b)–(d) Examples of the AITAH algorithm training. The training was done by manual annotation. Only the areas within the regions of interest (ROIs) shown as solid black lines were considered in the training. (b) original image, (c) manually annotated cells, (d) The AITAH algorithm training result. CD138-cells were marked cyan, and CD138+ cells were marked dark blue. (e)–(g) The training validation of the AITAH algorithm. The validation regions were marked as black dotted lines. (e) Analyzed images by the AITAH algorithm (CD138-cells marked cyan, CD138+ cells marked dark blue), (f) the validation from validator 1 (CD138-cells marked green, CD138+ cells marked yellow), (g) the validation from validator 2 (CD138-cells marked pink, CD138+ cells marked purple).

### AITAH algorithm validation and analysis

The accuracy and reliability of the AITAH algorithm were assessed through a two-step validation by the three external validators (O.L., J.K., and H.M.) who did not take part in the AITAH algorithm training to avoid biased decisions.

The training validation was performed in 18 WSIs not used for the model training. A total of 76 ROIs (36 regional and 40 object layers) were selected in each cycle phase and study group. The validators (O.L. and J.K.) annotated all the compartments, and the annotation was compared to the AITAH algorithm (**Fig 2e-g**). The error indices were automatically calculated in the AI platform^17^. Briefly, the total error indicates a sum of false negative (FN) and false positive (FP) predictions, precision is measured by dividing the true positive (TP) by all positives, sensitivity is measured by dividing the TP by the sum of the TP and the FN, and the F1 score is measured by the harmonic mean of precision and sensitivity. Furthermore, specificity is determined by dividing the true negative (TN) by the sum of TN and FP, while accuracy is computed by dividing the sum of TP and TN by the sum of TP, FP, FN, and TN.

In the performance validation, the validators (O.L., J.K., and H.M.) manually counted the number of stromal CD138+ cells in ten randomly selected high-power fields (HPFs) on each slide for a total of 10 slides. Since a 1-mm^2^ unit area is equal to four HPFs^29^, the cell counts from AITAH analysis were divided by 4 to convert to cells/HPF format. These converted data were compared to the median number of CD138+ cells determined by the validators.

After the validation, the analysis for 182 WSI (i.e., 71 control, 82 PCOS, and 29 RIF samples) was performed. CD138+ cell percentages were calculated manually by dividing the total number of CD138+ cells by the sum of CD138- and CD138+ cells in all ROIs within a WSI, and then multiplying by 100.

### Statistical analysis

Statistical analyses were conducted using IBM SPSS Statistics v28 (IBM Corp., Armonk, NY, United States), and visualization was created using GraphPad Prism (version 9.3.0) and RStudio (version 2022.12.0.353 with R version 4.2.2). For the women who had provided more than one sample, the CD138+ cell percentages and clinical features were counted as individual samples in each cycle phase. The cell percentages based on PCOS diagnosis and cycle phases were analyzed by the mixed- model analysis of variance (ANOVA), in which the total variance is divided into the within-group variance and the between-group variance. The continuous variables were tested by a Mann-Whitney U test for paired tests and a Kruskal-Wallis test for multiple comparisons depending on the data distribution. Statistical significance was defined as *p*<0.05. An intra-class correlation (ICC) estimate was calculated using a two-way mixed effects model with an absolute agreement model, with ICC values interpreted as poor (<0.5), moderate (0.5–0.75), good (0.75–0.9), and excellent (>0.9) reliability^30^. Correlations between the CD138+ plasma cell percentages and clinical characteristics were calculated using Spearman correlations, considering data availability and distribution.

## Results

### Training and validating the AI-algorithm AITAH

The final training error for the regional layers (i.e., epithelium and stroma) and the object layers (i.e., CD138-/+ stromal cell) was 2.28% and 6.15% (CD138+ cells 3.23%, CD138-cells 6.32%), respectively (**Supplemental Table S2**). In the training validation, median values of precision, sensitivity, F1 score, and specificity were calculated from 18 WSIs between pathologists (**Supplemental Figure S1a**). All verification statistics for CD138+ cells were 100%, indicating that the decisions made by the pathologists and the AITAH algorithm were in complete agreement. For the performance validation, we compared the number of CD138+ cells quantified by the pathologists and the AITAH analysis in 10 slides (**Supplemental Figure S1b**). The two evaluation methods showed excellent accuracy (ICC; 0.76, 95% CI; 0.36 to 0.93, *p*=0.002) and a positive correlation (Spearman’s rank correlation coefficient: 0.79, *p*<0.01). The interobserver variability between pathologists indicated good to excellent reliability for training validation and good reliability for performance validation (**Supplemental Table S3**). Therefore, the AITAH algorithm demonstrated sufficient functionality in identifying the target plasma cells.

### Endometrial stromal CD138+ plasma cell percentages by cycle phase and PCOS status

The AITAH analyzed 71 controls and 82 PCOS slide images, grouped by PCOS diagnosis/phenotypes and menstrual cycle phases. CD138+ cell percentages showed a significant decrease from proliferative (PE) to secretory phases, regardless of PCOS diagnosis (*p*<0.001) (**Fig 3a**). Moreover, we did not find any differences in the CD138+ cell proportion between control and PCOS cases at the same cycle phase (*p*_PE_=0.83, *p*_ESE_=0.22, *p*_MSE_=0.92, *p*_LSE_=0.98). Interestingly, anovulatory PCOS cases exhibited similar CD138+ cell percentages than the PCOS SE samples (*p*>0.58) but markedly lower than PCOS PE samples (*p*<0.001). In the phenotype comparisons, we pooled the three secretory phases into one phase (SE) to simplify the comparisons between the cycle phases and increase the power of statistical analysis. Phenotype A cases showed significantly higher CD138+ cell percentages compared to phenotype D cases in the PE (*p*<0.001) (**Fig 3b**).

**Figure 3.**
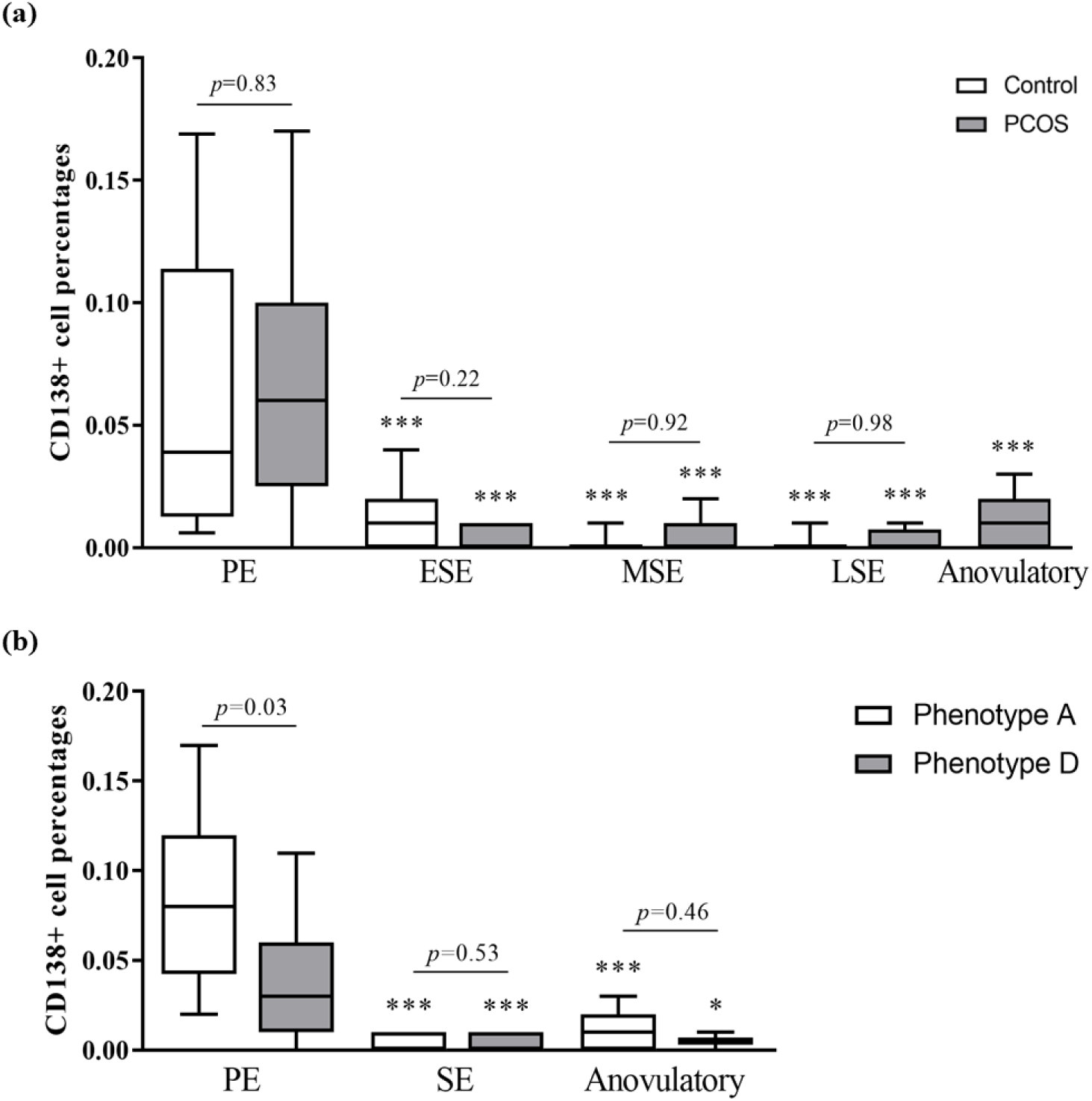
**Stromal CD138+ plasma cell percentages across PCOS status and cycle phases.** Stromal CD138+ cell percentages for (a) controls (12 PE, 15 ESE, 25 MSE, 19 LSE) and PCOS cases (21 PE, 14 ESE, 21 MSE, 16 LSE, 11 Anovulatory) and (b) different PCOS phenotypes (PhenotypeA; 12 PE, 26 SE, 8 Anovulatory, Phenotype D; 9 PE, 16 SE, 2 Anovulatory). Phenotype C was excluded due to the small sample size. The box indicates the inter-quartile range, the middle line represents the median, and the whiskers show the min-max range. \*\*\**p*<0.001 when compared to the PE. The statistical differences were calculated by the mixed model ANOVA. PE (proliferative phase), ESE (early secretory phase), MSE (mid-secretory phase), LSE (late secretory phase), SE (secretory phase)

### Correlations between endometrial stromal CD138+ plasma cell percentages and clinical characteristics in women with PCOS and non-PCOS controls

To validate the correspondence between the AITAH analysis and menstrual cycle-related traits and elucidate factors contributing to increased CD138+ cell percentages in the PE, we analyzed correlations between the plasma cell percentages and physiological features (**Table 2**). Control SE samples showed negative correlations with P4 (r^2^=-0.43, *p*=0.02) and positive correlations with FAI and LH (FAI; r^2^=0.45, *p*<0.001, LH; r^2^=0.41, *p*=0.02). In contrast, PCOS SE cases exhibited positive correlations with testosterone (r^2^=0.32, *p*=0.02) and AMH (r^2^=0.30, *p*=0.03). Meanwhile, we did not observe any significant correlations in the PCOS anovulatory group.

**Table 2.** Correlations between the CD138+ cell percentages and endometrium thickness and hormonal values.

|  |  | CD138+<br>percentages | Endometrial<br>thickness (mm) | P4<br>(nmol/L) | Testosterone<br>(nmol/L) | FAI | FSH<br>(IU/L) | LH<br>(IU/L) | AMH<br>(ng/ml) |
| --- | --- | --- | --- | --- | --- | --- | --- | --- | --- |
| **Control** |  |  |  |  |  |  |  |  |  |
| Control<br>(n=12) | PE | 0.04<br>[0.01,0.11] | 5.25<br>[4.43,5.78]<br>(-0.16) | 14.60<br>[1.34,33.34]<br>(-0.035) | 1.01<br>[0.80,1.38]<br>(0.06) | 2.20<br>[1.50,2.76]<br>(0.13) | 7.41<br>[6.68,8.75]<br>(0.33) | 7.69<br>[6.70,9.23]<br>(0.19) | 2.28<br>[1.22,4.64]<br>(-0.40) |
| Control<br>(n=59) | SE | 0.00<br>[0.00,0.01] | 9.80<br>[8.40,11.30]<br>(0.21) | 25.35<br>[4.78,41.61]<br>**(-0.43\*\*)** | 1.17<br>[0.95,1.65]<br>(-0.11) | 2.00<br>[1.24,2.93]<br>**(0.45\*\*\*)** | 3.68<br>[2.61,4.69]<br>(0.15) | 7.64<br>[4.82,10.34]<br>**(0.41\*\*)** | 2.25<br>[1.35,4.48]<br>(0.06) |
| **PCOS** |  |  |  |  |  |  |  |  |  |
| PCOS<br>(n=21) | PE | 0.06<br>[0.03,0.10] | 5.35<br>[4.53,6.03]<br>(0.02) | 11.21<br>[0.25,48.12]<br>(0.33) | 1.00<br>[0.83,1.77]<br>(-0.02) | 2.64<br>[1.87,3.22]<br>(0.30) | 6.79<br>[6.13,8.63]<br>**(-0.52\*)** | 9.72<br>[8.47,11.27]<br>(-0.03) | 5.27<br>[2.67,7.40]<br>(0.24) |
| PCOS<br>(n=50) | SE | 0.00<br>[0.00,0.01] | 9.70<br>[8.00,10.50]<br>(-0.05) | 22.49<br>[0.37,41.06]<br>(-0.24) | 1.33<br>[1.00,1.68]<br>**(0.32\*)** | 2.51<br>[1.61,3.34]<br>(0.09) | 3.57<br>[2.62,5.03]<br>(-0.03) | 8.97<br>[5.19,13.20]<br>(0.14) | 3.68<br>[2.50,5.83]<br>**(0.30\*)** |
| PCOS<br>Anovulatory<br>(n=11) |  | 0.01<br>[0.00,0.02] | 5.60<br>[5.00,8.95]<br>(0.05) | 0.29<br>[0.26,0.33]<br>(-0.17) | 1.65<br>[1.24,2.49]<br>(-0.47) | 6.42<br>[3.34,11.82]<br>(-0.05) | 6.59<br>[5.65,7.59]<br>(0.36) | 16.46<br>[10.69,20.05]<br>(0.03) | 9.08<br>[5.01,12.80]<br>(0.02) |

The CD138 cell percentages and clinical characteristics presented as median with interquartile range [Q1;Q3]. The correlations are presented in brackets. *p* values were determined using log-transformed values of sex-hormone-related characteristics for the skewed data distribution. \* *p*<0.05, \*\* *p*<0.01, and \*\*\* *p*<0.001
P4 (progesterone), FAI (free androgen index), FSH (follicle-stimulating hormone), LH (luteinizing hormone), AMH (anti-Müllerian hormone)

### Endometrial stromal CD138+ plasma cell percentages by receptivity in RIF samples

Next, we investigated potential variations in CD138+ cell percentages across distinct endometrial receptivity statuses, defined by the gene expression. We utilized the endometrial samples that were timed after progesterone administration at P4+5 days, which is considered the window of implantation and common time for embryo transfer in assisted reproductive technology. We did not observe any differences in CD138+ cell percentages between different receptivity statuses among RIF samples (*p*=0.81) (**Fig 4**).

**Figure 4.**
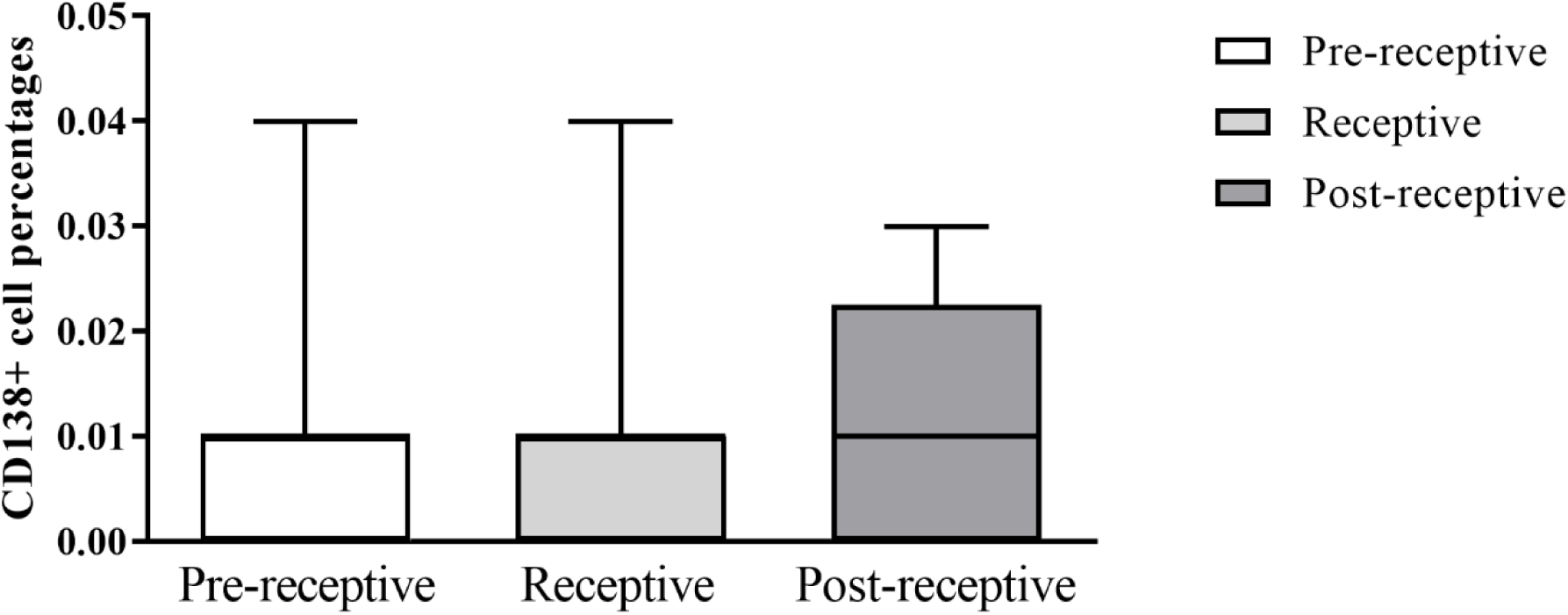
**Stromal CD138+ plasma cell percentages stratified by endometrial receptivity.** The CD138+ cell percentage comparisons of RIF patient samples (9 RIF pre-receptive, 10 RIF receptive, 10 RIF post-receptive). The box indicates the inter-quartile range, the middle line represents the median, and the whiskers show the min-max range. The statistical differences were calculated by the Kruskal-Wallis test.

## Discussion

In this study, we developed an AI algorithm, AITAH, for the first time, to quantify CD138+ plasma cells and investigate the factors influencing the CD138+ cell aggregation in the human endometrium without any other signs of CE. We verified that the AITAH algorithm demonstrated excellent precision and specificity compared to human analysis. Our findings align with the previous research showing a higher CD138+ plasma cell presence in the proliferative phase endometrium compared to the secretory phase endometrium^8,31^. As a novel finding, anovulatory PCOS samples showed significantly lower CD138+ cell percentages compared to the PE PCOS samples, and phenotype A PCOS samples showed higher CD138+ cell percentages in the proliferative phase than phenotype D ones. On the other hand, the cell percentages did not show variations based on endometrial receptivity in RIF samples.

In recent years, the advancement of deep CNN technology has revolutionized the rapid analysis of a massive number of WSIs. However, the application of CNN algorithms in uterine studies has predominantly focused on cancer research^18,32^. To our knowledge, our study was the first to target endometrial CD138+ plasma cells in a whole view of slides using a cloud-based CNN platform with high precision and specificity. As supported by previous publications, the implementation of the CNN platform enhances analysis capabilities by reducing intra- and inter-observer variations, minimizing hands-on time, and improving the speed and reproducibility of analysis^15,17,25^. Indeed, we analyzed 182 WSIs within an hour, surpassing what could be achieved by a pathologist alone. Still, certain endometrial features posed challenges in AITAH algorithm training and analysis, particularly pre- decidual structures in the stroma. AITAH occasionally misidentified these structures as epithelium, thus ignoring the stromal cells in those areas. Nevertheless, based on previous studies using the same AI platform, we evaluated the overall performance to be excellent ^14,15,17^.

Higher CD138+ cell percentages in the proliferative phase compared to the secretory phase may be due to the estradiol-mediated recruitment of plasma cell progenitors from the systemic circulation into the endometrial mucosa, supported by a mouse study^33^. Furthermore, as the endometrium thickens towards the secretory phase, biopsies may predominantly contain superficial layers, resulting in lower CD138+ cell percentages relative to unstained stromal cells^31^. These hypotheses do not, however, explain our results that there were fewer CD138+ cells in anovulatory and amenorrhoeic PCOS samples compared to proliferative PCOS samples, despite the similar endometrial thickness. Considering that anovulatory endometrium exhibits an increased expression of estrogen receptors, heightened estrogen sensitivity, and greater estrogen exposure^19,34^, further research with a larger sample size is necessary to explore the presence of CD138+ cells in the anovulatory PCOS endometrium. In addition, higher CD138+ percentages in PCOS phenotype A compared to D at the beginning of a menstrual cycle (cycle days 6-8) suggest that hyperandrogenism also affects CD138+ recruitment.

Our result indicated that receptivity status at P4+5 had no effect on CD138+ cell accumulation. However, further investigation is warranted to validate previous findings suggesting an increased prevalence of endometrial inflammation (or even CE) in infertile women as assessed by CD138+ plasma cell aggregation^7,8^ and the potential detrimental impact of HRT on immune cell profiles^35^. Future studies should consider larger sample sizes and incorporate prospective study designs, including women undergoing IVF treatment with natural menstrual cycles or stimulated cycles versus HRT cycles.

The main strength of our study is the reliability of our AI algorithm in identifying stromal CD138+ plasma cells within a WSI in only a few seconds. Furthermore, with the rare and well-characterized human tissue sample set, we were able to encompass diverse endometrial conditions: i) different menstrual cycle phases; ii) individuals with PCOS or RIF diagnoses; iii) both ovulatory and anovulatory cases; and iv) varying endometrial receptivity statuses.

As for limitations, we lacked confirmed CE cases as positive controls, although we did not look for overt endometritis. Additionally, the limited number of anovulatory PCOS samples may explain the absence of correlations between CD138+ plasma cell percentages and clinical characteristics, despite the exceptionally large total PCOS sample set. We lacked detailed clinical information on the RIF patients such as serum sample analysis. Lastly, in both control and PCOS populations, serum estrogen levels fell below the detection limit by LC-MS, as all steroids were analyzed in a single run and unavailable for separate analysis. Despite these limitations, our study showed the promising potential of AI technology to address current clinical and practical challenges in detecting endometrial immune cells.

## Conclusion

Our findings emphasize the accuracy and potential of AITAH algorithm in detecting endometrial CD138+ plasma cells, offering distinct advantages such as rapid inspection of whole slide images, independence from trained pathologists, and consistent productivity. This supports the utilization of AI technology to help clinical decision-making for example in endometrial cycle phase-related dynamics, as well as in different reproductive disorders.

## Supplemental Data

**Supplemental Figure S1. The AITAH algorithm validation result.** Validation for the AITAH algorithm was carried out in two stages: (A) training validation and (B) performance validation. (A) The heatmap calculated by the median between the two validators presents the agreements between the AITAH algorithm and the validators. (B) The number of CD138+ cells per high power field (HPF) from manual counting was compared to the AITAH analysis results. The area unit (mm^2^) was converted to HPF (1 HPF=0.25 mm^2^). The validation values were calculated by the median between the three validators.

**Supplemental Table S1. Baseline characteristics of PCOS subjects by phenotypes**

**Supplemental Table S2. The AITAH algorithm training results**

**Supplemental Table S3. Interobserver variability in the AITAH algorithm validation**

## Ethics Statement

This study was approved by The Regional Ethics Committee of the Northern Ostrobothnia Hospital District, Finland (65/2017), and the Research Ethics Committee of the University of Tartu, Estonia (340T-12). All experiments were performed in accordance with relevant guidelines/regulations in line with the Declaration of Helsinki.

## Author Contributions

T.T.P., A.S., R.K.A., and S.L designed the study. T.T.P., A.S., R.K.A., E.K.K., and M.S. contributed to sample collection. A.A., K.K., and M.S. were involved in sample processing. O.L., J.A.K., and H.M. performed the validation. S.L. and E.K.K. performed image analysis. S.L. and R.K.A. performed the statistical analysis. All authors revised and approved the final version.

## Acknowledgments

This study would not have been possible without the participation of all of the subjects. We thank research nurse Elina Huikari for her help in sample collection, and Riitta Vuento for her technical assistance in the IHC staining.

## Data availability statement

The datasets generated during and/or analysed during the current study are not publicly available due to sensitivity of the health data. Non-personal data can be requested from the corresponding author.

