## Supplemental Table/Figure for "AI-algorithm training and validation for endometrial CD138+ cells in infertility-associated conditions; polycystic ovary syndrome (PCOS) and recurrent implantation failure (RIF)"

### Supporting information

**Supplemental Figure 1. The AITAH algorithm validation result.**

**Supplemental Figure 2. Stromal CD138+ plasma cell percentages depending on PCOS phenotype**

**Supplemental Table 1. The AITAH algorithm training results**

**Supplemental Table 2. Interobserver variability in the AITAH algorithm validation**

**Supplemental Figure 1. The AITAH algorithm validation result.**

Validation for the AITAH algorithm was carried out in two stages: (A) training validation and (B) performance validation. (A) The heatmap calculated by the median between the two validators presents the agreements between the AITAH algorithm and the validators. (B) The number of CD138+ cells per high power field (HPF) from manual counting was compared to the AITAH analysis results. The area unit (mm2) was converted to HPF (1 HPF=0.25 mm2). The validation values were calculated by the median between the three validators.

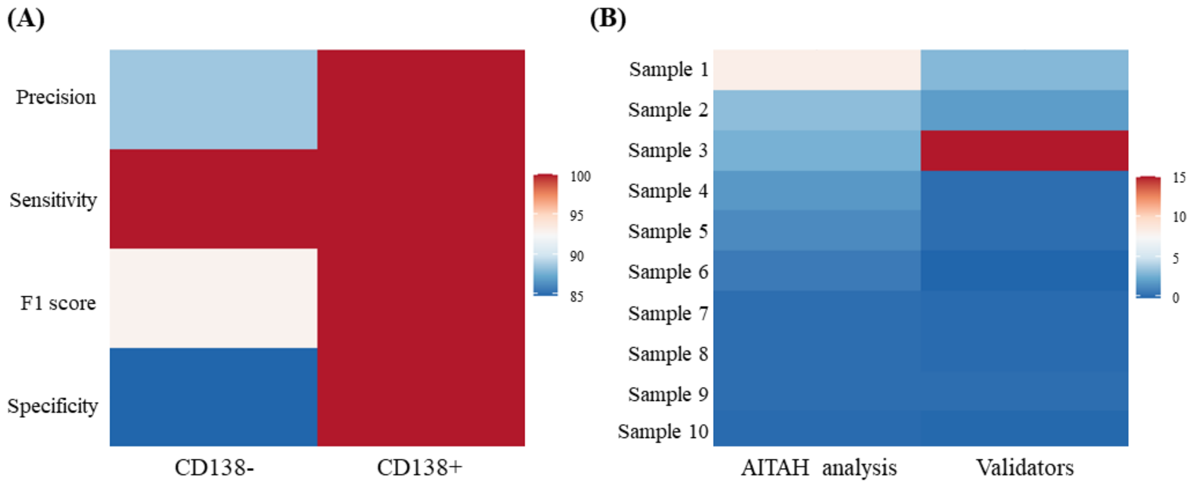

**Supplemental Figure 2. Stromal CD138+ plasma cell percentages depending on PCOS phenotype**

Stromal CD138+ cell percentages in different PCOS phenotypes; PCOM+OA+HA (Phenotype A; 9 PE, 26 SE, 7 Anovulatory) and PCOM+OA (Phenotype D; 8 PE, 16 SE, 2 Anovulatory). PCOM+HA group (Phenotype C; 6 SE) was excluded as the sample size was small and the samples were collected at SE only. ***p*<0.01, and ****p*<0.001 by Kruskal-Wallis test. The box indicates the inter-quartile range, the middle line represents the median, and the whiskers show the min-max range.

OA (oligo-anovulation), HA (hyperandrogenism), PCOM (polycystic ovaries), PE (proliferative phase), SE (secretory phase)

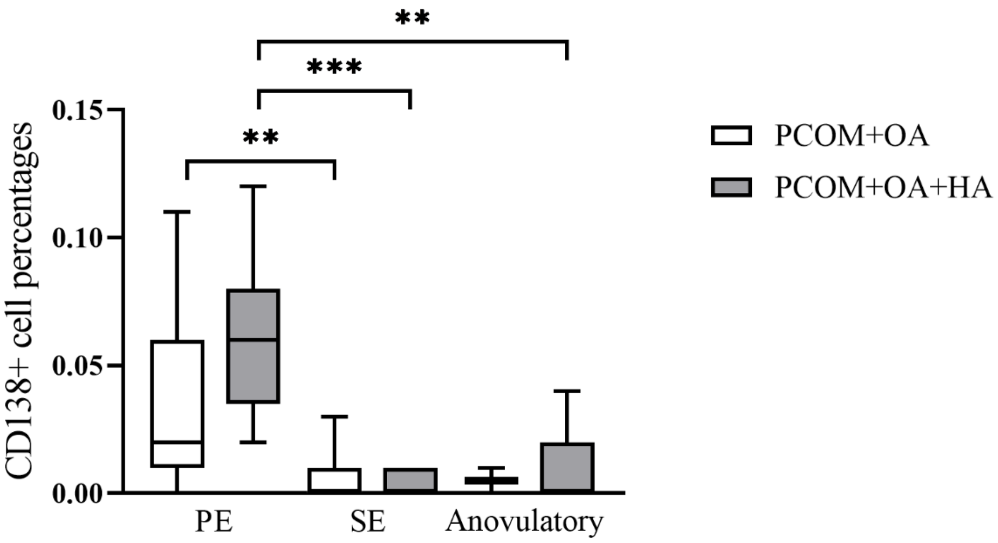

**Supplemental Table 1. The AITAH algorithm training results**

| (%) | CNN 1 | Epitehlium | Stroma | CNN 2 | CD138+ | CD138- |
|---|---|---|---|---|---|---|
| Total error | 2.28 | 15.20 | 1.04 | 6.15 | 3.23 | 6.32 |
| Precision | 99.17 | 92.86 | 99.82 | 99.45 | 100 | 99.42 |
| Sensitivity | 98.46 | 91.86 | 99.14 | 94.36 | 96.77 | 94.22 |
| F1 score | 98.81 | 92.36 | 99.48 | 96.84 | 98.36 | 96.75 |
| Specificity | 99.60 | 92.94 | 99.82 | 99.48 | 100 | 99.45 |
| Accuracy | 99.23 | 92.40 | 99.23 | 96.92 | 98.39 | 96.84 |

CNN 1 consists of epithelium and stroma, and CNN2 consists of CD138+ and CD138- cells. The F1 score was measured by the harmonic mean of precision and sensitivity. CNN (convolutional neural network)

**S2 Table. Interobserver variability in the AITAH algorithm validation**

| ICC (95% CI) | Epithelium | Stroma | CD138- cell | CD138+ cell |
|---|---|---|---|---|
| Training<br>between two pathologists | 0.933***<br>(0.869,0.966) | 0.927***<br>(0.855,0.963) | 0.858***<br>(0.719,0.927) | - |
| Performance<br>between three pathologists | | | | 0.820**<br>(0.486,0.951) |

* *p*<0.05, ** *p* <0.01, *** *p* <0.001

ICC (intraclass correlation coefficient), CI (confidence interval)
